# Engaging communities through participatory learning action for the control and prevention of diabetes: a protocol for the Process Evaluation of the EMPOWER-D trial in Pakistan and Afghanistan

**DOI:** 10.64898/2026.03.05.26347686

**Authors:** Maria Ishaq Khattak, Khalid Rehman, Saima Afaq, Sabeen Saeed Butt, Gul Ghutai, Refat Hanifi, Murtaza Hofiani, Amber Tahir, Rubia Zafar, Hannah Maria Jennings

## Abstract

**Background:** Type 2 diabetes is a growing challenge in low- and middle-income countries, where health systems face major capacity gaps. Participatory learning and action (PLA) has shown effectiveness in preventing type 2 diabetes in Bangladesh, but little is known about its use in other LMICs for diabetes. The EMPOWER-D (Engagement of community through Participatory learning and action for cOntrol and prevention of type 2 diabetes) trial is testing PLA for diabetes prevention in communities in Pakistan and Afghanistan. This protocol describes the plans for the embedded process evaluation (PE).

**Methods:** The PE will use a mixed-methods design across three sites, following the UK Medical Research Council framework for PE, examining implementation, mechanisms of impact and context. Implementation will be assessed using adaptation reports, fidelity checklists, attendance data, and supervisor reports. Mechanisms of impact will be explored through interviews, focus group discussions and photovoice. Contextual factors will be examined through interviews with participants, community mobilisers, supervisors, and key stakeholders. Quantitative data will be analysed descriptively, while qualitative data will undergo thematic analysis using a theory of change framework. Comparative analysis will identify common and context-specific influences.

**Discussion:** This is the first multi-country PE of a PLA intervention for diabetes prevention to our knowledge, and the first in Afghanistan and Pakistan. The study will provide insights into how the intervention was delivered, how and why it worked (or did not work), and the contextual factors shaping outcomes. Findings will inform the adaptation and scale-up of participatory approaches for non-communicable disease prevention in resource strained setting health systems. Trial registration: ClinicalTrials.gov: NCT06561126 (registered 23 August 2024); NCT06570057 (registered 26 August 2024).

## Introduction

Pakistan and Afghanistan are facing an escalating public health crisis posed by type 2 diabetes mellitus (T2DM) [1, 2]. According to the International Diabetes Federation Pakistan has the highest global age-standardised prevalence of T2DM, with 31.4% of adults aged 20-79 years (approximately 34.5 million individuals) affected in 2024 [3]. An estimated 26.9% of these cases are undiagnosed, equating to 9.28 million individuals [3]. The number of people living with diabetes in Pakistan is projected to rise to 70.2 million by 2050, which a 103% increase [2, 3]. Afghanistan also shows concerning trends, with an age-standardised diabetes prevalence of 11.7% affecting 1.93 million adults in 2024, projected to rise to 4.76 million by 2050, with 71.4% undiagnosed [4].

The growing burden of T2DM in both countries is driven by a complex interplay of genetic, environmental, and behavioural factors [5, 6]. Evidence from LMICs shows that non-pharmacological and behavioural interventions can prevent or delay the onset of T2DM and its complications [7]. Community-based interventions are a particularly effective strategy in preventing T2DM [8]. Participatory Learning Action (PLA) is an approach to research and interventions where communities are empowered to identify and address their own problems and solutions [9]. It has been systematised into a groups-based intervention where community groups regularly meet and follow a cycle of: identifying problems, planning solutions to those problems, implementing solutions and reflecting on them [10]. Originally developed to improve maternal and neonatal health outcomes in LMICs where it was highly effective [11, 12], this community-based PLA was later adapted to address diabetes prevention and control in Bangladesh through the DMAGIC (Community groups or mobile phone messaging to prevent and control type 2 diabetes and intermediate hyperglycaemia in Bangladesh) project, and it was tested through a cluster-randomised controlled trial and found to be effective at preventing T2DM at a population level [10, 13].

### The EMPOWER-D project

The effectiveness of the DMAGIC PLA intervention in rural Bangladesh [14, 15] provides a strong rationale for its adaptation to neighbouring countries [16]. The “Engagement of Community through Participatory Learning and Action for Control and Prevention of T2DM and its Risk Factors” (EMPOWER-D) project will adapt, implement, and test this community-based PLA intervention to prevent and manage T2DM and its risk factors in Pakistan and Afghanistan through three interrelated studies:

i. Rural Pakistan (Khyber Pakhtunkhwa): A cluster randomised controlled trial in two sites (Peshawar and Swabi) to test the effectiveness of the intervention at a population level (ClinicalTrials.gov ID: NCT06561126).
ii. Urban Pakistan (Karachi, Sindh): A feasibility cluster randomised controlled trial will assess the potential for conducting a full-scale trial to examine the impact of the intervention in urban settings of Karachi (ClinicalTrials.gov ID: NCT06570057).
iii. Rural Afghanistan (Greater Kabul region): A feasibility trial to evaluate the feasibility of implementing and testing the intervention in a conflict-ridden rural setting such as Afghanistan.

At each site, the trial will be conducted across clusters that are randomly assigned to intervention or control arms. Baseline and endline assessments will be conducted at all three sites, and data will be collected through surveys, anthropometric measurements, and blood tests. In parallel with these trials, mixed-method process evaluations (PE), described in this manuscript, will be conducted to examine how and why the intervention works, or fails to work, across different contexts. PEs can help explain how and why interventions work (or do not) providing important insights that cannot be captured in outcome evaluations [17–19]. PEs are considered particularly valuable for complex interventions, where mechanisms of change and contextual factors including culture, gender dynamics, health system capacity, and local resources are understood to strongly shape outcomes [19–22]. While the EMPOWER-D intervention is being implemented in three distinct contexts and each site is adapting it accordingly (publication forthcoming), it is guided by a shared theoretical framework. The PE will enable us to compare across sites how it is implemented and its mechanisms.

### The EMPOWER-D intervention and theory of change

The PLA approach draws on Paulo Freire’s concept of “critical consciousness.” Freire proposed that when marginalised communities critically reflect on their circumstances and engage in collective dialogue, they become empowered to take actions that lead to positive social change [10, 23, 24]. Within this framework, knowledge is co-produced, and community members are positioned as active agents rather than passive recipients of health information [25]. Evidence from DMAGIC demonstrated that the intervention worked by both enabling individuals to change their behaviours and by shifting community-level environments and norms, including improvements in knowledge and social practices [10, 26].

Drawing on the philosophy of PLA, the planned intervention and trial design, the socio-ecological model, contextual knowledge, and findings from the DMAGIC process evaluation, the research team developed a theory of change (ToC) for the adapted EMPOWER-D intervention (Figure 1). This process was undertaken collaboratively, with input from representatives across all study sites. We acknowledge there will be differences across sites – for example in Afghanistan, women’s movement is particularly restrictive. The ToC proposes that through the formation of women’s and men’s groups, participants will strengthen their knowledge, expand their social networks, and enhance collective capacity. These mechanisms are expected to contribute to improved mental and physical health and enable individuals, families, and communities to address the causes and consequences of diabetes.

**Figure 1.**
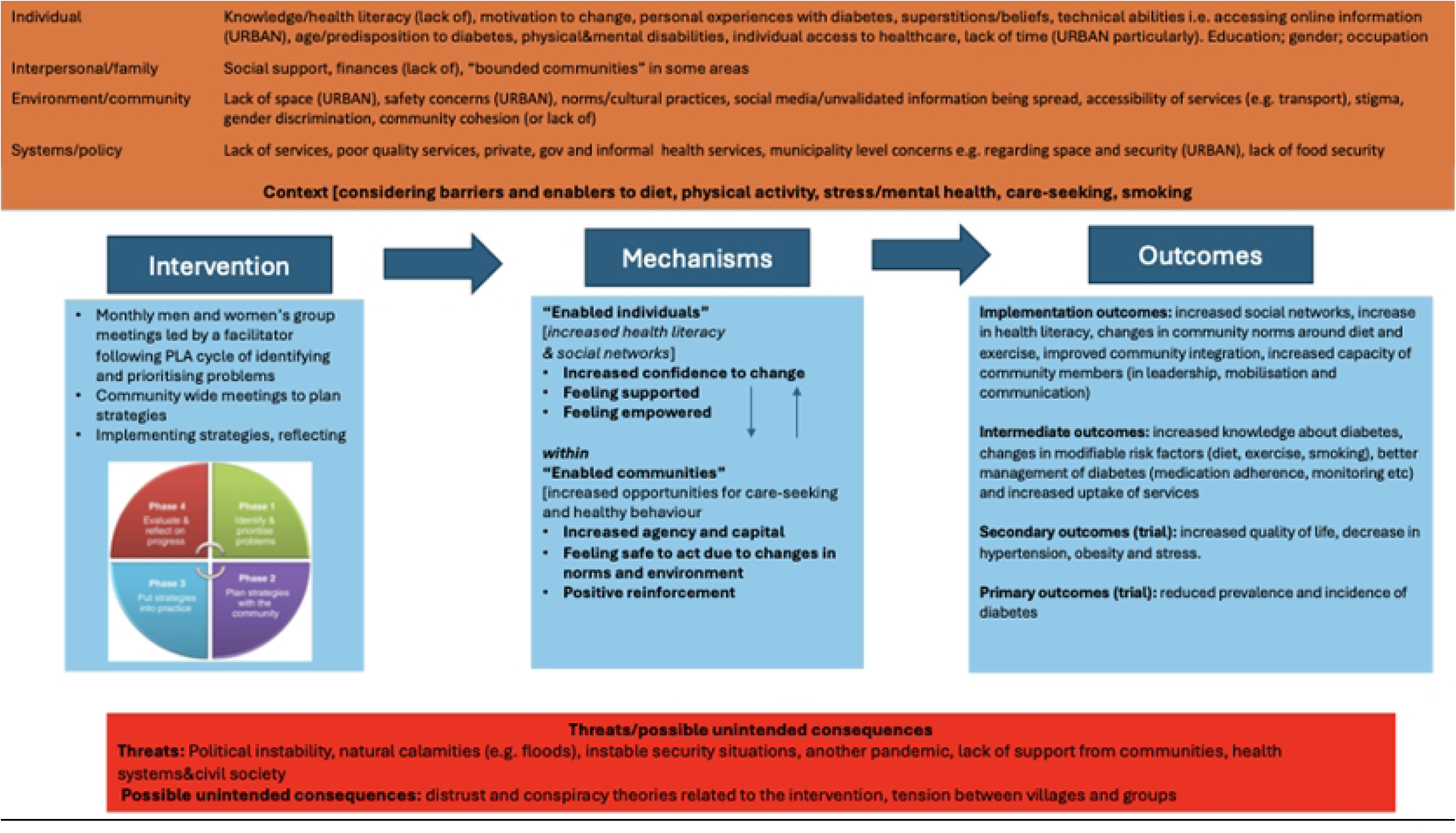
EMPOWER-D working theory of change

The EMPOWER-D intervention, similar to the DMAGIC intervention, involves translating the PLA philosophy into practice through facilitated men and women’s group meetings in which communities identify priority health problems, plan strategies collectively, implement chosen solutions, and reflect on their effectiveness (figure 2) [10]. Community mobilisers (CM) will be recruited from the local community to facilitate group meetings. CMs will be equipped with a structured manual, an illustrated flipbook, and a toolkit of participatory activities to guide the process. Group meetings should be interactive and dialogical, promoting active participation and encouraging members to share learning and implement strategies between sessions. The PLA cycle consists of both village level group meetings and wider community level meetings. The cycle will be 18-months in Pakistan, and has been condensed to 12-months for Afghanistan.

**Figure 2.**
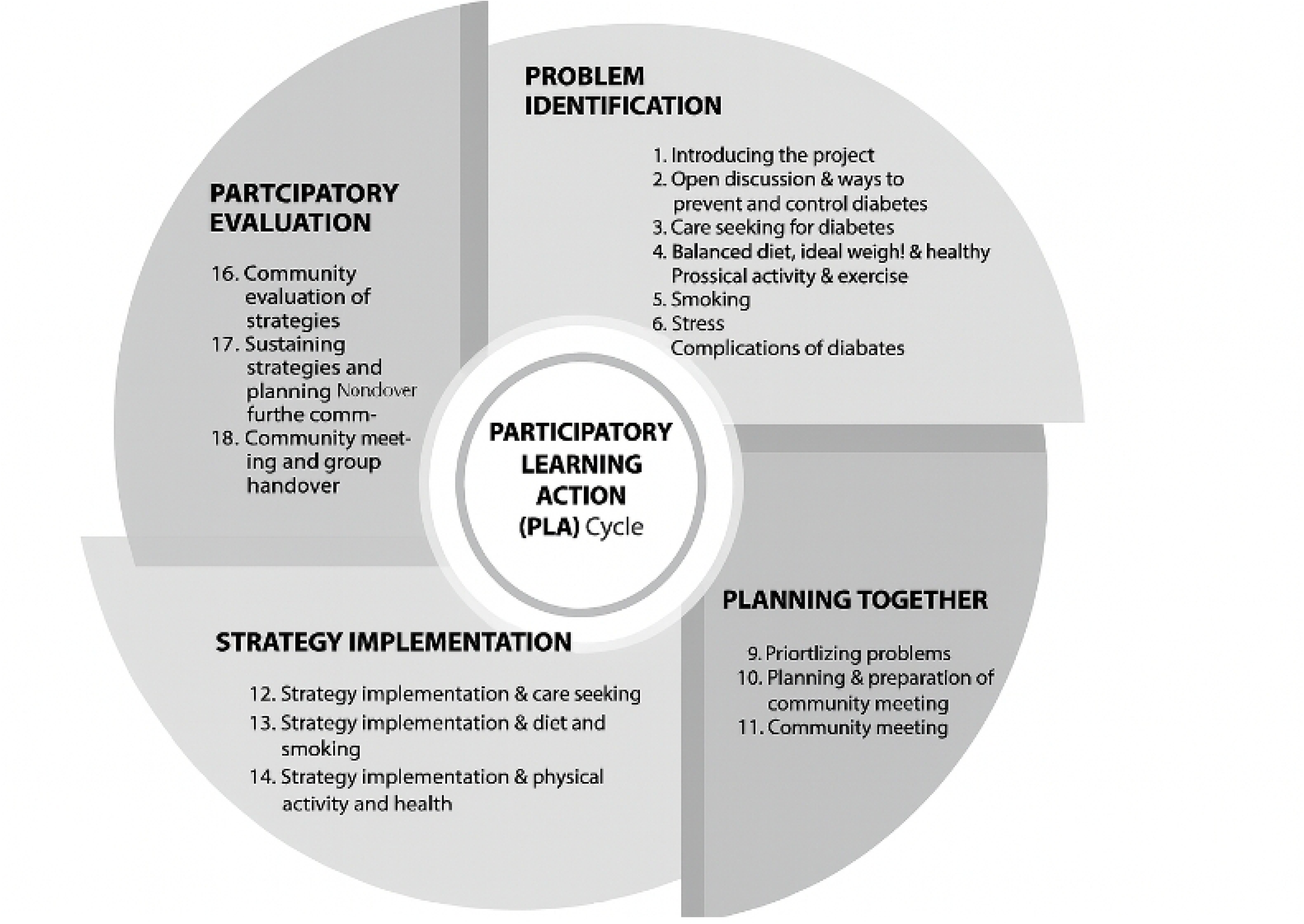
Participatory learning and action cycle and group meetings, figure from Morrison et al. [10]

The aim of this PE is “*to explore how the EMPOWER-D intervention is implemented across different sites, what works and leads to change, what does not work, and how context affects the intervention.*”

## Methods

### Overall approach

The PE will follow the MRC framework for process evaluations of complex interventions (Figure 3). This involves examining three key domains:

i. Implementation: This includes the implementation process and what was delivered. We will examine: adaptations made, fidelity, dose, and reach. We will also examine the acceptability of the intervention and the potential for scale-up and implementation beyond the trial period.
ii. Mechanisms of impact: It looks at how the intervention worked or did not work. We will examine participant responses to the intervention, mediators (drawing on our ToC), and any unexpected consequences of the intervention.
iii. Context: This refers to factors (cultural, social, economic etc.) external to the intervention that may influence the intervention’s delivery, how it is received as well as how context influences any changes in behaviour related to diabetes more generally.

**Figure 3.**
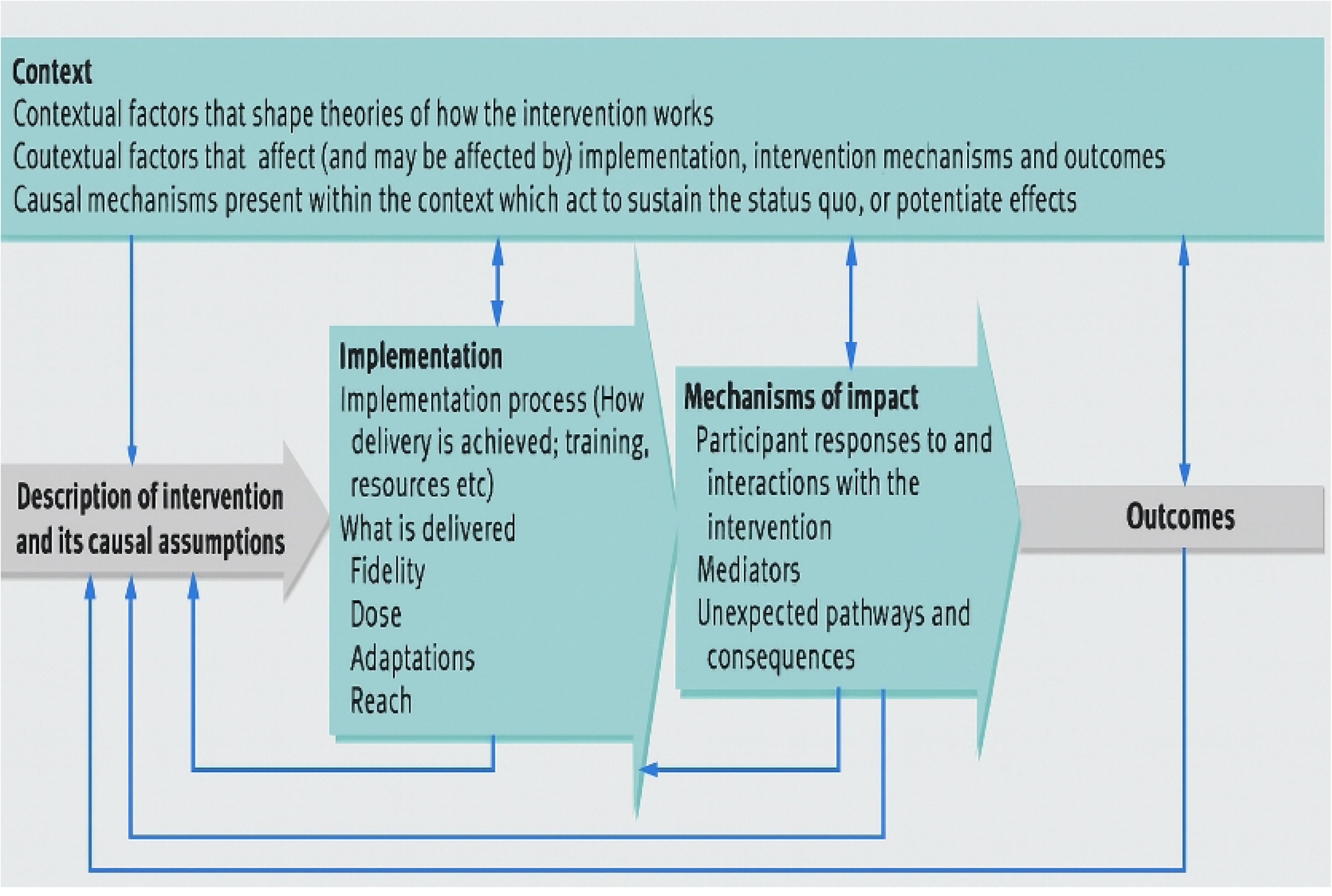
Medical Research Council framework for process evaluation of complex interventions, figure from Moore et al., 2018 [18]

We will employ a convergent mixed-methods design, integrating both qualitative and quantitative approaches across all sites. The methods will include in-depth interviews (IDIs), focus group discussions (FGDs), photovoice, and quantitative data drawn from project reports, attendance registers, supervisor checklists, monitoring forms, and trial survey datasets. Additionally monthly/quarterly reports will be reviewed for insights. For each domain different methods will be used, and initially quantitative and qualitative data will be analysed separately. The results will be integrated according to the domain/component of the PE, location and planned outputs. Table 1 outlines the planned methods, analyses, and expected outputs for each domain and component of the PE. By employing a diverse set of methods, this evaluation will generate rich and triangulated insights, ensuring both breadth and depth in understanding the implementation, mechanisms of action, and contextual factors. Detailed descriptions of each method are provided in the subsequent section (data collection).

**Table 1.**
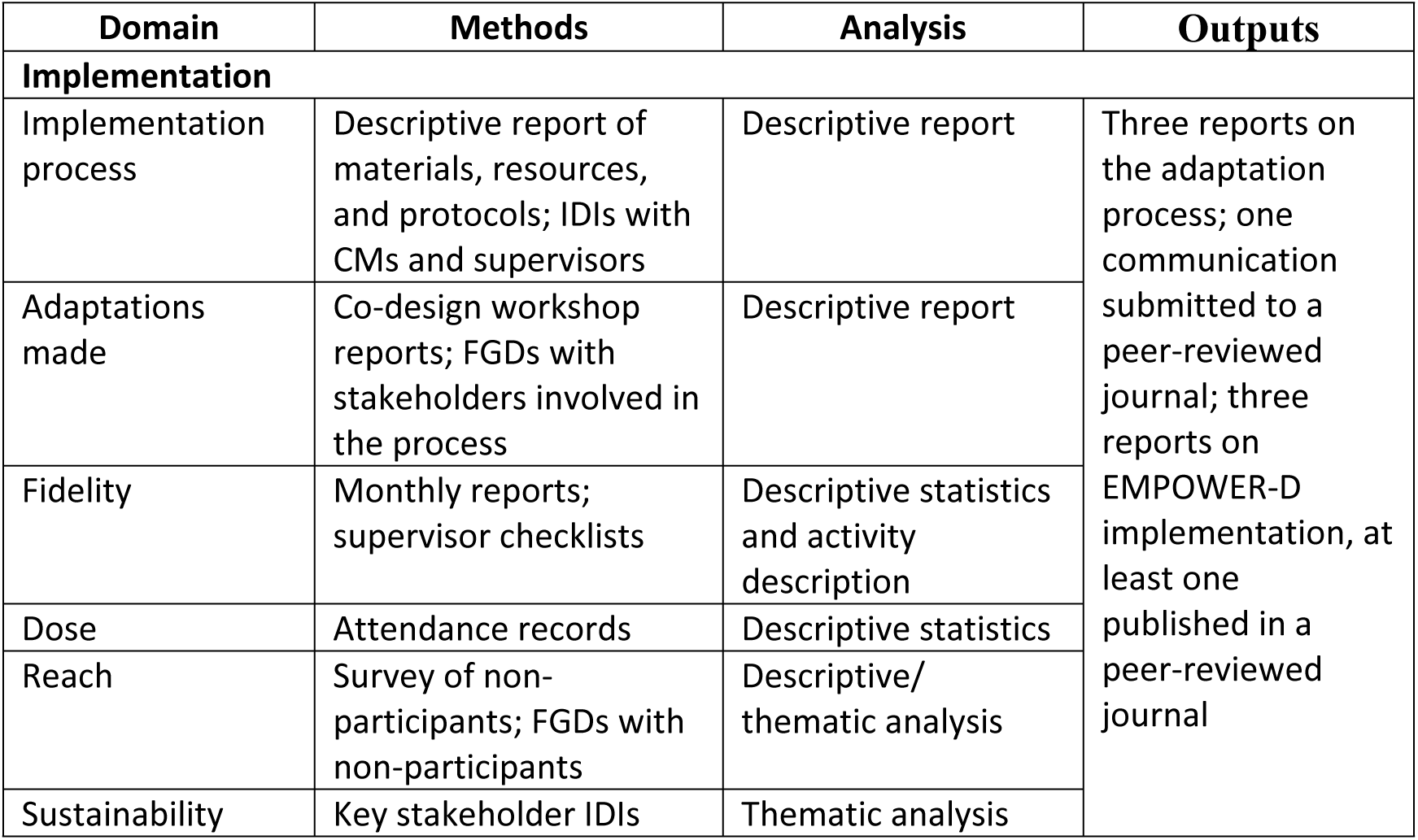
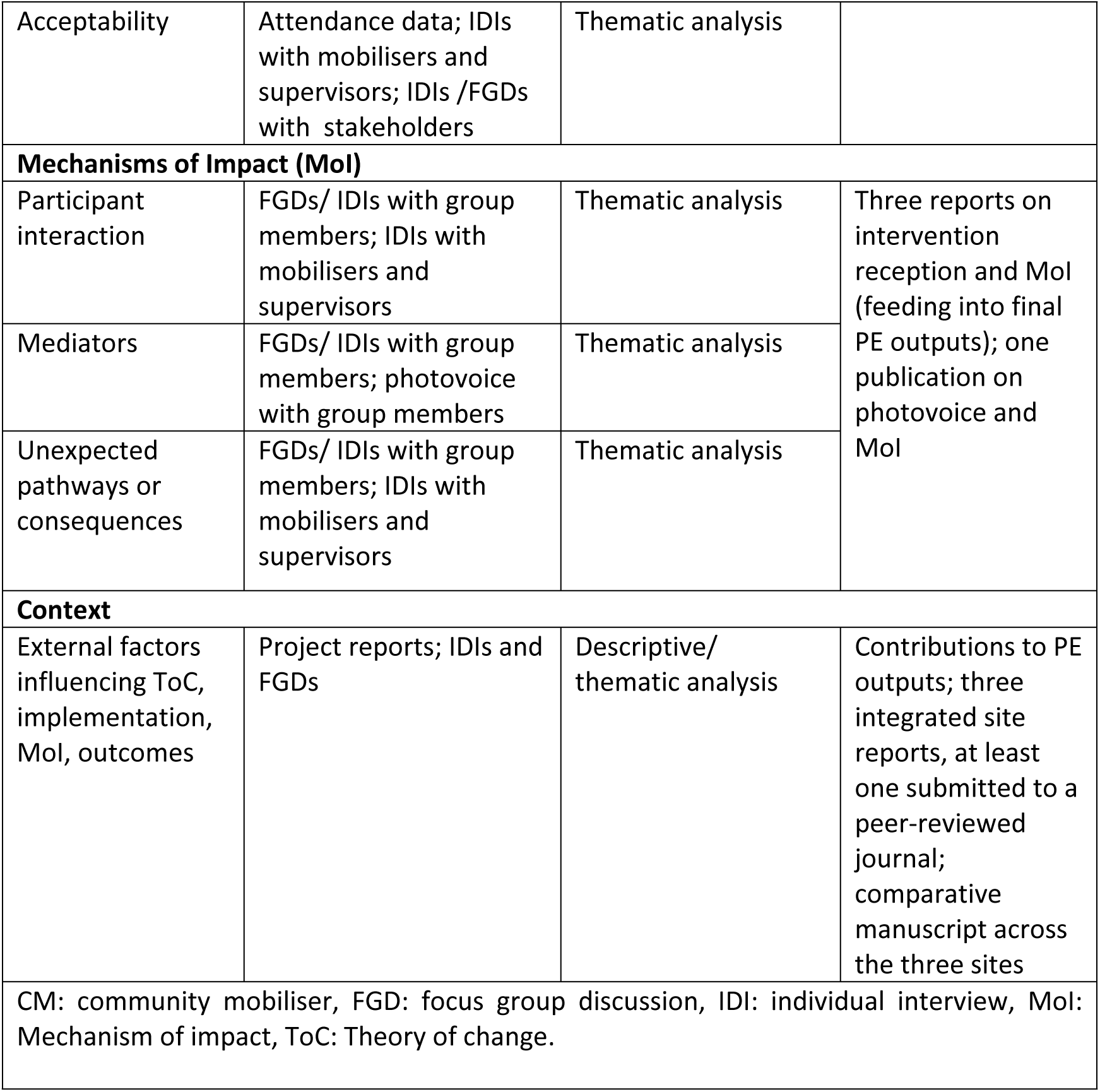
Analytical framework for process evaluation.

| Domain | Methods | Analysis | Outputs |
| --- | --- | --- | --- |
| **Implementation** |  |  |  |
| Implementation process | Descriptive report of materials, resources, and protocols; IDIs with CMs and supervisors | Descriptive report | Three reports on the adaptation process; one communication submitted to a peer-reviewed journal; three reports on EMPOWER-D implementation, at least one published in a peer-reviewed journal |
| Adaptations made | Co-design workshop reports; FGDs with stakeholders involved in the process | Descriptive report |  |
| Fidelity | Monthly reports; supervisor checklists | Descriptive statistics and activity description |  |
| Dose | Attendance records | Descriptive statistics |  |
| Reach | Survey of non-participants; FGDs with non-participants | Descriptive/thematic analysis |  |
| Sustainability | Key stakeholder IDIs | Thematic analysis |  |
| Acceptability | Attendance data; IDIs with mobilisers and supervisors; IDIs /FGDs with stakeholders | Thematic analysis |  |
| **Mechanisms of Impact (Mol)** |  |  |  |
| Participant interaction | FGDs/ IDIs with group members; IDIs with mobilisers and supervisors | Thematic analysis | Three reports on intervention reception and Mol (feeding into final PE outputs); one publication on photovoice and Mol |
| Mediators | FGDs/ IDIs with group members; photovoice with group members | Thematic analysis |  |
| Unexpected pathways or consequences | FGDs/ IDIs with group members; IDIs with mobilisers and supervisors | Thematic analysis |  |
| **Context** |  |  |  |
| External factors influencing ToC, implementation, Mol, outcomes | Project reports; IDIs and FGDs | Descriptive/ thematic analysis | Contributions to PE outputs; three integrated site reports, at least one submitted to a peer-reviewed journal; comparative manuscript across the three sites |
| CM: community mobiliser, FGD: focus group discussion, IDI: individual interview, Mol: Mechanism of impact, ToC: Theory of change. |  |  |  |

### Data collection according to PE domain

#### Implementation

##### Adaptations

Adaptations made to the intervention from each site followed a systematic process of formative research and a series of co-design workshops (publication forthcoming) – the research teams produced reports documenting the process, key decisions, and final adaptations. In addition, the PE team will conduct FGDs with those involved in the adaptation of the intervention including: researchers, workshop participants and community advisory panel (CAP) members. The FGDs will explore the adaptation process, contextual influences, challenges encountered, facilitators, and lessons learned.

##### Implementation process

At each site the training materials and protocols, monthly supervisor reports and checklists, and project reports will be systematically reviewed. Each site will generate a descriptive report of the implementation process, highlighting activities undertaken, variations across villages or groups, stakeholders involved, and associated costs and time commitments. These reports will be complemented by IDIs with CMs and supervisors to identify barriers and facilitators influencing implementation.

##### Fidelity

Fidelity refers to the degree to which the intervention is delivered as intended. In this study, fidelity will be assessed across three dimensions: (1) adherence of group meetings to the overall PLA process; (2) the extent to which CMs followed the intervention manual, including key facilitation behaviours such as encouraging participation, asking open-ended questions, and fostering group engagement; and (3) the participatory nature of the intervention, as reflected in the levels of involvement and engagement of both group members and the wider community. Fidelity will be evaluated using: (i) structured supervisory checklists completed during observations of monthly meetings; (ii) monthly reports from supervisors and data collectors documenting group activities; and (iii) qualitative IDIs with CMs and supervisors to capture their perspectives on fidelity and contextual variations across sites.

##### Dose and reach

In this evaluation, dose refers to the intensity of the intervention, assessed through indicators such as the number of participants attending group meetings, the proportion of community members engaged in the intervention, the frequency of participation, and the extent of involvement in group-led strategies and activities. Reach is defined as the extent to which the intervention influenced individuals beyond the immediate PLA groups, e.g. through knowledge dissemination and behavioural change within the wider community. Quantitative data will be drawn from attendance logs and monthly supervisor reports, which will capture information on the number and duration of group sessions, attendance patterns, and levels of participant engagement. Additionally, survey data will include items on attendance at regular group meetings and broader community sessions, familiarity with the PLA intervention and group activities, and any reported changes in knowledge, attitudes, or behaviours attributable to the intervention. To complement the quantitative data, qualitative methods will be used to explore perceptions of dose and reach. FGDs will be conducted with non-group members to understand their awareness, interaction with, and perceived impact of the intervention. Further, FGDs and IDIS with CMs, supervisors, and group members will be carried out to gather detailed insights into how intensively the intervention was delivered and how far its influence extended within the community.

##### Acceptability and sustainability

This component of the evaluation aims to understand how the intervention was received culturally and socially, and to assess the potential for its continuation and scale-up beyond the study period. Acceptability will be examined through FGDs and IDIs with group members. These discussions will focus on participants’ interactions with the intervention, their perceptions of its relevance and usefulness, the extent of its alignment with local cultural norms, and the barriers and facilitators encountered during implementation, with particular attention to their perspectives on the participatory approach and its compatibility with existing community structures. Additionally, IDIs will be conducted with community stakeholders (including healthcare workers, community leaders, and religious scholars) exploring views on the cultural and community relevance of the intervention, levels of community engagement, and perspectives on opportunities for long-term sustainability, integration into local health/civil society structures, and the feasibility of scaling up the intervention. Demographic data will be collected on group attendance to identify patterns of participation across gender, age, and socioeconomic groups. Where gaps are observed (e.g. underrepresentation of certain groups), these issues will be explored during FGDs to understand underlying reasons.

### Mechanisms of impact

To understand how the intervention worked (and did not work), drawing on the study’s ToC this domain of the PE will explore the three interrelated levels of change:

i. Individual level: changes in self-confidence, personal agency, empowerment, perceived safety, and adherence to healthier behavioural practices.
ii. Group level: dynamics of group interaction, development of collective efficacy, and peer support mechanisms.
iii. Community level: shifts in community norms, social cohesion, support for positive health behaviours, enabling or constraining factors for change, and observable transformations within the wider community over the course of the intervention.

We will also examine unintended consequences or challenges encountered during implementation.

IDIs and FGDs will be conducted with PLA group members, CMs, and facilitators. These will explore participants’ engagement in the intervention, experiences of behaviour change, perceived individual and collective empowerment, community level effects, and challenges or opportunities that emerged during implementation. Photovoice methodology will be employed specifically in rural Pakistan (KP) as part of FGDs with group members. Participants will be invited to document their experiences of the intervention using photographs, which they will bring to the FGDs. These images will serve as visual prompts for discussion, facilitating the expression of participants’ perspectives on the changes they experience and the challenges they face. FGDs will then be used to interpret the photographs and explore the broader meanings and social context embedded in these visual narratives.

### Context

Context will be examined to explore external factors that influence the implementation and outcomes of the intervention. They may include cultural, social, spatial, environmental, and political factors. In order to explore the impact of context, we will review the monthly project reports, and questions will be asked regarding the influence of context during the FGDs and IDIs with PLA group members, community members, and supervisors.

### Data analysis

Table 2 outlines all data collection methods, the analysis plan and how they will contribute to the planned outputs. There are three main types of data: qualitative (FGDs, IDIs, photovoice), project reports and quantitative. The data will initially be analysed separately. Quantitative data will initially be collected on ODK or Microsoft Excel. It will be analysed using Microsoft and Stata to generate descriptive statistics, capturing relevant domains of the evaluation (reach, fidelity, dose) and enabling comparisons over time. The interviews and FGDs will be recorded and transcribed verbatim and translated into English. Documents and qualitative data will be analysed thematically following a process of: immersion in data, coding data, grouping codes into themes, reviewing and defining themes and writing up findings [27, 28]. Coding and data organisation will be supported using ATLAS.ti and/or NVIVO [29, 30], which will aid in the systematic identification of themes, subthemes, and cross-cutting patterns within and across sites. The analysis will be guided by both inductive and deductive approaches, allowing for the emergence of context-specific insights while also testing assumptions derived from the intervention’s ToC. Particular attention will be given to how implementation processes, contextual conditions, and participant engagement influence mechanisms of impact. Qualitative and quantitative data will be integrated at the interpretation and reporting stage – according to the paper and output (see table 2). Triangulation of the data will strengthen the validity and depth of conclusions. This mixed-methods strategy will enable a comprehensive assessment of how and why the EMPOWER-D intervention was implemented, what outcomes it generated, and under what conditions these outcomes were realised. Full details of data sources, timelines, and analytical responsibilities are presented in Table 2.

**Table 2:**
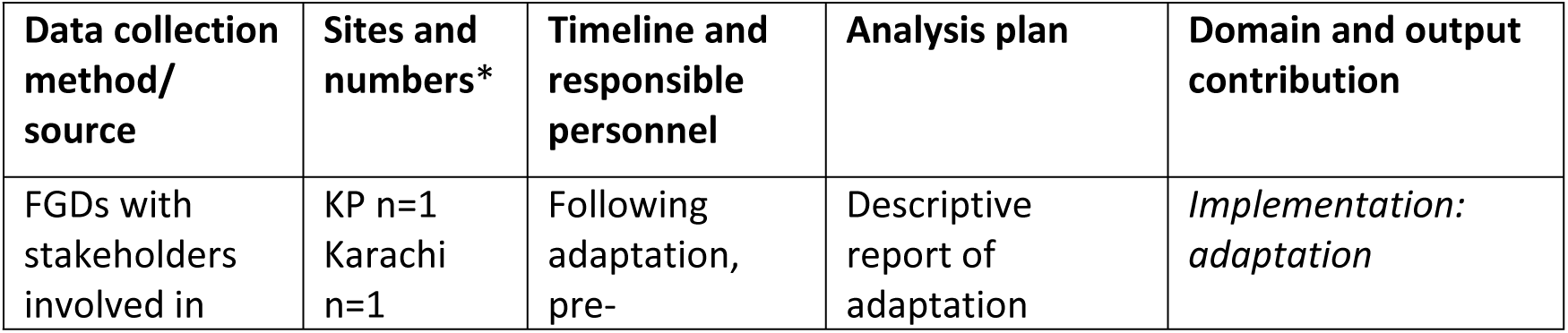
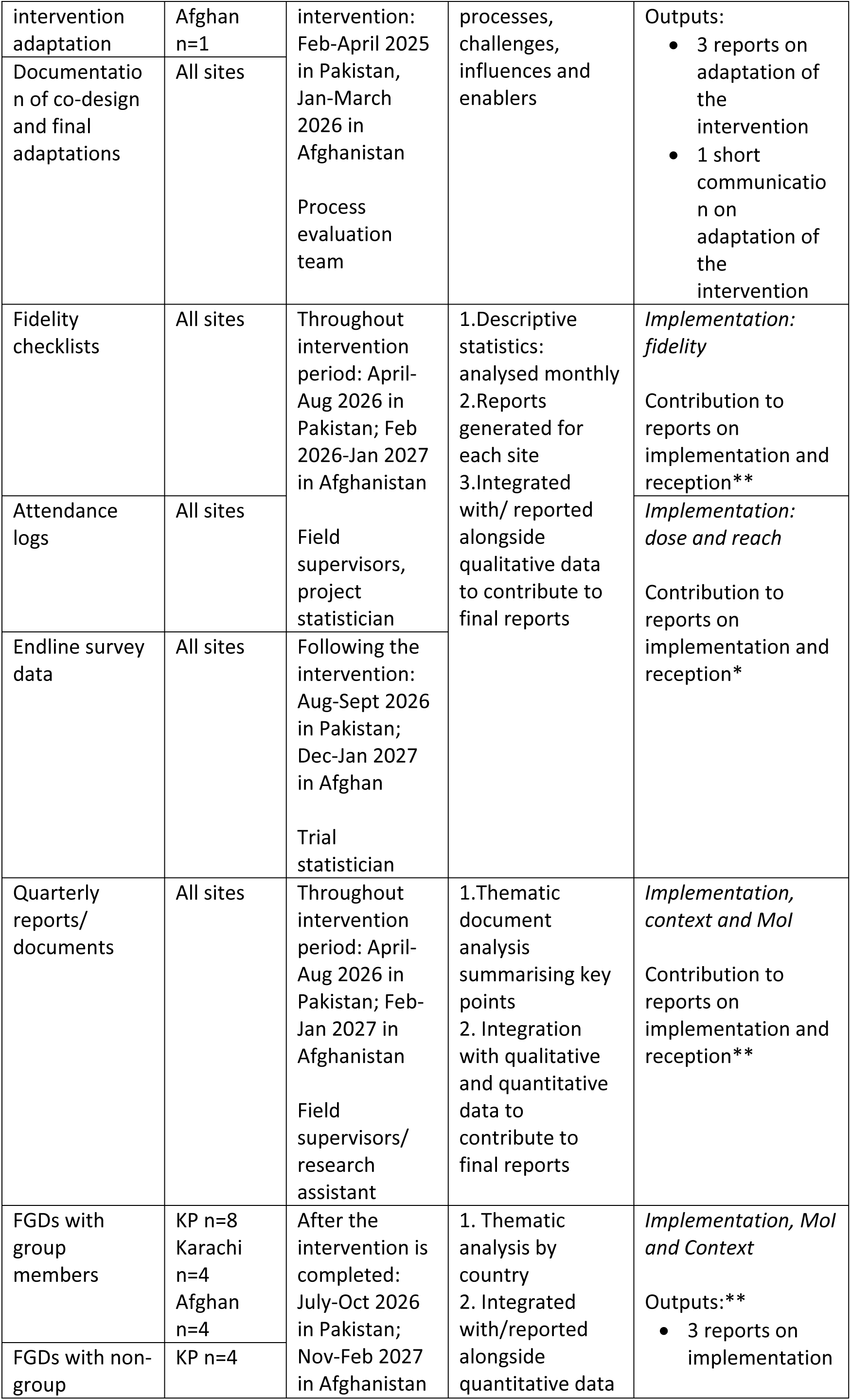
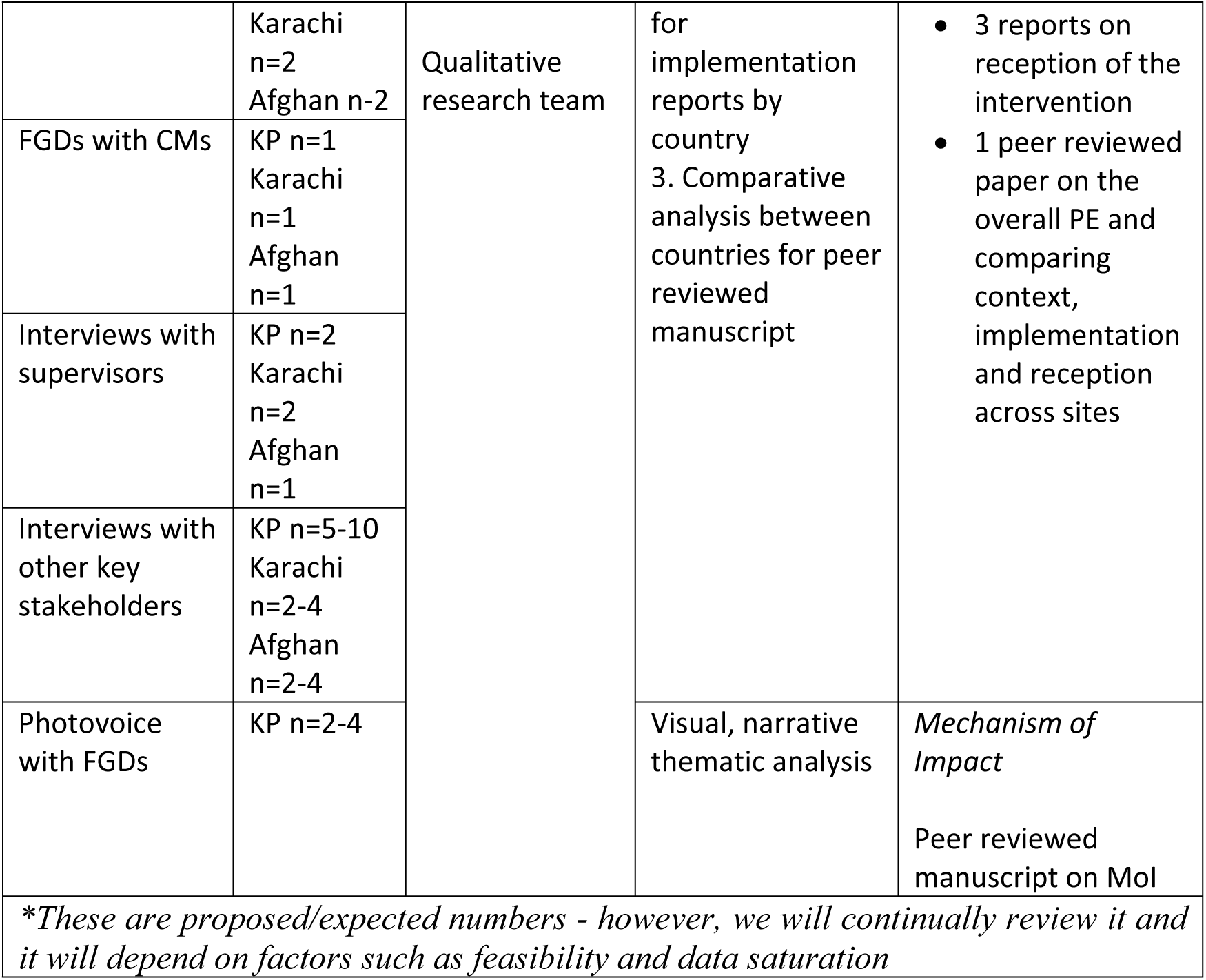
Data collection and analysis plan.

#### Overall timeline of the study

The intervention began in March 2024 in Pakistan and February 2025 in Afghanistan, and is due to complete September 2026 in Pakistan and January 2027 in Afghanistan. Data collection began pre-intervention (for reporting of the adaptation) in January 2024. Most data collection will continue throughout the intervention period (regular reporting, checklists and logs) and last until after the intervention (FGDs, IDIs) which will be October 2026 for Pakistan and February 2027 for Afghanistan. Participants who participated in the intervention and trial will be recruited for interviews and FGDs (begun in January 2025) – with initial recruitment for qualitative data being in July 2026 in Pakistan and December 2026 in Afghanistan, and complete by September 2026 for Pakistan and February 2027 for Afghanistan. We expect analysis to be completed and results shared by mid-2027. Specific details for the timeline for each aspect of the study are in table 2.

### Quality assurance and data management

To ensure the rigour, reliability, and ethical integrity of the process evaluation, comprehensive quality assurance and data management procedures will be implemented across all sites. These procedures are designed in line with best practices for conducting mixed-methods research and process evaluations of complex interventions [10, 31, 32].

All data collection tools, including topic guides, supervisory checklists, and survey questionnaires, will be standardised across sites and pilot-tested prior to implementation. Field teams will receive thorough training on research protocols, ethical conduct, and context-specific considerations. Training will include instruction on obtaining informed consent, maintaining confidentiality, and the use of data collection software and devices [33].

Quantitative data will be collected using digital platforms where feasible and managed using Stata [34]. Data entry procedures will include validation rules and regular cleaning routines to minimise errors [35]. Descriptive statistical analyses will be performed for supervisory checklists, attendance data, and survey responses. Qualitative data will be audio-recorded (with participant consent), transcribed ad verbatim, translated into English where necessary, and anonymised using unique participant identifiers. Transcripts will be securely stored in encrypted folders, accessible only to authorised research team members. Data will be managed using ATLAS.ti software and/or NVIVO to facilitate systematic coding and analysis [28, 36]. Coding reliability will be enhanced through regular team coding meetings and inter-coder reliability checks [37].

Regular quality control mechanisms will include spot checks by supervisors, weekly debriefing meetings, and monthly reviews of field data [38]. These checks will enable early identification and resolution of inconsistencies or protocol deviations [39]. A centralised data coordination team will oversee data harmonisation across sites and support integration for cross-site analysis. These quality assurance practices aim to enhance the validity, credibility, and transferability of the findings [40], ensuring that the process evaluation provides robust evidence on the implementation, mechanisms of action, and contextual factors shaping the EMPOWER-D intervention.

### Ethics

This process evaluation is part of the approved EMPOWER-D trial protocol and has received ethical clearance from the National Bioethics Committee of Pakistan (Approval No. 4-87/NBCR-1070/23/2026) and the Aga Khan University Ethics Review Committee (Approval No. 2025-9340-34977). All participants will be informed about the purpose of the evaluation, the voluntary nature of their participation, and their right to withdraw at any time. Written informed consent will be obtained from all interview and FGD participants. Anonymity and confidentiality will be maintained through pseudonymisation of transcripts and secure storage of audio files and data. Only authorised members of the research team will have access to the data.

### Dissemination plans

Findings from this PE will be disseminated through a range of channels. Dissemination activities will include presentations at national and international conferences, preparation and distribution of policy briefs to implementing partners and decision-makers, and submission of manuscripts to peer-reviewed journals.

## Discussion

This study is among the first multi-country PE of a community-based PLA intervention for T2DM in LMICs, and the first to include a fragile setting such as Afghanistan. Guided by the MRC framework, it aims to generate evidence on the delivery of the EMPOWER-D intervention, the mechanisms through which it operates, and the contextual factors shaping outcomes across Pakistan and Afghanistan. Publishing this protocol enhances transparency and contributes to strengthening the reporting of process evaluations in LMICs.

This PE study addresses several gaps in the literature. It will provide insight into understanding how community engagement and a participatory community intervention may work or not work in this context, and how in practice it can be implemented, offering important insights into any future work. There have been few PEs conducted across multiple LMIC sites, and none have systematically included fragile contexts [41]. Creative participatory approaches remain under-utilised in LMIC health research, as shown by a scoping review of 57 studies in which participatory systems mapping predominantly engaged policymakers and facilitated learning but rarely included formal evaluation or produced actionable outcomes [42].

Despite a growing recognition of the importance of process evaluations, there remains relatively limited evidence on the utilisation of PE of complex interventions from LMICs where financing, infrastructure, and workforce capacity are constrained [43]. This highlights the importance of formative engagement with local stakeholders to ensure interventions are responsive, contextually appropriate, and adaptable to fragile health systems. The EMPOWER D PE uses a ToC to link activities with outcomes at individual, group, and community levels, testing assumptions about change and showing how cultural norms, gender relations, and health systems shape results, with a mixed methods design using IDIs, FGDs, monitoring data, and photovoice to ensure triangulation and a comprehensive account of implementation.

Conducting a PE across three sites will present challenges, as variations in research capacity may affect data quality, security risks in Afghanistan and cultural norms may restrict participation, particularly for women and marginalised groups, and attrition may reduce representation of those who disengage from the intervention; these risks will be addressed through rigorous training, purposive sampling, and continuous quality assurance.

In summary, the EMPOWER D process evaluation will generate novel evidence on the implementation and adaptation of PLA interventions for diabetes prevention in LMIC and fragile settings, enrich interpretation of trial outcomes, advance methodological learning on process evaluation, and provide transferable lessons for scaling participatory approaches to address the rising burden of NCDs in resource constrained health systems.

## Data Availability

No datasets were generated or analysed during the current study. All relevant data from this study will be made available upon study completion.

## Acknowledgements

We are very grateful to the adaptation and intervention team of EMPOWER-D, particularly Dr Noor Sanauddin and Dr Inayat Shah, for their contributions to the initial discussions and shaping of the protocol during the early phases of the project. We also thank the community mobilisers, field supervisors, and members of the Community Advisory Panels in Pakistan and Afghanistan for their valuable support in guiding this process evaluation.

